# Two years after lockdown: longitudinal trajectories of sleep disturbances and mental health over the COVID-19 pandemic and the effects of age, gender, and chronotype

**DOI:** 10.1101/2022.07.29.22278180

**Authors:** Federico Salfi, Giulia Amicucci, Domenico Corigliano, Lorenzo Viselli, Aurora D’Atri, Daniela Tempesta, Maurizio Gorgoni, Serena Scarpelli, Valentina Alfonsi, Michele Ferrara

## Abstract

Since the first lockdown of Spring 2020, the COVID-19 contagion waves pervasively disrupted the sleep and mental health of the worldwide population. Notwithstanding the largest vaccination campaign in human history, the pandemic has continued to impact the everyday life of the general population for two years now. The present study provides the first evidence of the longitudinal trajectories of sleep disturbances and mental health throughout the pandemic in Italy, also describing the differential time course of age groups, genders, and chronotypes.

A total of 1062 Italians participated in a three-time points longitudinal study covering two critical stages of the emergency [the first lockdown (April 2020) and the second lockdown (December 2020)] and providing a long-term overview two years after the pandemic outbreak (April 2022). We administered validated questionnaires to evaluate sleep quality/habits, insomnia, depression, stress, and anxiety symptoms.

Analyses showed a gradual improvement in sleep disturbances, depression, and anxiety. Conversely, sleep duration progressively decreased, particularly in evening-type and younger people. Participants reported substantial earlier bedtime and get-up time. Stress levels increased during December 2020 and then stabilised. This effect was stronger in the population groups apparently more resilient during the first lockdown (older people, men, and morning-types).

Our results describe a promising scenario two years after the pandemic onset. However, the improvements were relatively small, the perceived stress increased, and the re-establishment of pre-existing social/working dynamics led to general sleep curtailment. Further long-term monitoring is required to claim the end of the COVID-19 emergency on Italians’ sleep and mental health.

## Introduction

In the first months of 2020, the severe acute respiratory syndrome coronavirus 2 (SARS-CoV-2) started to spread worldwide, giving rise to a pandemic. To deal with the increasing contagion and death rates due to virus propagation, governments around the world started to apply extraordinary containment measures consisting of home confinement, social distancing, and the closure of most business activities. The lockdown period was associated with raised sleep disturbance and mental health problems among the general population, as reported by consistent meta-analytic literature (Jahrami et al., 2021, 2022; Robinson et al., 2022). Italy was the first European country to handle the contagion wave of COVID-19, implementing a total lockdown lasting two months (March–April 2020). Several Italian studies confirmed the pervasive impact of this unprecedented period on sleep quality/habits and psychological well-being (Casagrande et al., 2020; Cellini et al., 2020; Salfi, Lauriola, et al., 2021). After a phase of alleviated pandemic emergency and loosened restrictions (Summer 2020), a second large contagion wave occurred in Autumn 2020. The Italian government promptly reacted to the new exacerbated scenario by applying partial lockdown measures on regional basis, weighted according to the local load on the healthcare system and infection rates.

Notwithstanding the adoption of this regional approach consisting of lighter restraining measures than the first lockdown, some longitudinal investigations showed a persistent impact of the emergency period on sleep (Conte et al., 2021; Salfi, D’Atri, et al., 2021) and mental health (Salfi, D’Atri, et al., 2021) among the Italian population during the second contagion wave. These results were consistent with the international literature confirming long-lasting repercussions on sleep features (Basishvili et al., 2021; Liu et al., 2022; Trakada et al., 2022) and psychological measures (Benke et al., 2022; Chodkiewicz et al., 2021; Daly & Robinson, 2022; Rus Prelog et al., 2022; Wetherall et al., 2022) during the second COVID-19 wave, suggesting the urgency of large-scale interventions to preserve the general well-being. In the last days of 2020, the administration of the first COVID-19 vaccine to the adult population was authorised by the European and Italian medicines agencies. Notwithstanding the largest vaccination campaign in human history and the consequent lifting of restraining measures in the subsequent months, the COVID-19 has continued to disrupt the everyday life of the worldwide population for two years now. However, the long-term impact of the pandemic on sleep and mental health remains poorly elucidated. In this vein, the first aim of the present study is to identify the longitudinal trajectories of sleep quality, insomnia, depression, stress, and anxiety in the general population across the pandemic in Italy. We surveyed a large sample of Italian citizens (N = 1.062) using validated questionnaires at three time points: during the first weeks of lockdown (April 2020), during the second contagion wave (December 2020), and two years after the first implementation of the lockdown measures (April 2022).

The current literature consistently showed that age groups (Amicucci et al., 2021; Bottary et al., 2022; Daly et al., 2020; Jahrami et al., 2022; Rossi et al., 2020), genders (Daly et al., 2020; Rossi et al., 2020; Salfi et al., 2020; Salfi, Lauriola, et al., 2021), and chronotypes (Bottary et al., 2022; Merikanto et al., 2022; Salfi, Lauriola, et al., 2021) reacted differently to the first months of restraining measures. Specifically, younger people (Amicucci et al., 2021; Daly et al., 2020; Jahrami et al., 2022; Rossi et al., 2020) and women (Daly et al., 2020; Rossi et al., 2020; Salfi et al., 2020; Salfi, Lauriola, et al., 2021) reported higher rates of sleep disturbances and psychological symptoms. These results were confirmed by Italian (Salfi, D’Atri, et al., 2021) and European studies (Benke et al., 2022; Chodkiewicz et al., 2021; Rus Prelog et al., 2022; Wetherall et al., 2022) addressing the effect of the second wave of COVID-19. Similarly, the evening chronotype was associated with more evident changes in sleep patterns and increased sleep and mental health problems both during the lockdown (Bottary et al., 2022; Merikanto et al., 2022; Salfi, Lauriola, et al., 2021) and the second contagion wave (Salfi, D’Atri, et al., 2021). Based on this evidence, another objective of the present research is to identify the different time courses of sleep and psychological disturbances between age groups, genders, and chronotypes. Our longitudinal investigation is unique in covering two critical stages of the COVID-19 outbreak (the first lockdown and the second partial lockdown) and providing a long-term overview of the Italians’ general well-being two years after implementing the first measures to counteract the virus’ spread.

## Methods

### Participants and procedure

The present study consists of a longitudinal web-based survey involving three assessment points. Participants responded using an online platform (*Google forms*). The first survey wave was held during the third and fourth weeks of the lockdown period of Spring 2020 (25 March–7 April 2020), coinciding with the first contagion peak of COVID-19. Subsequently, respondents were invited to participate in the second survey wave by email during the contagion peak of the second pandemic wave (28 November–11 December 2020). Finally, all respondents were re-invited to take part in the third survey wave two years after the first one (9 April–22 April 2022). A total of 1062 Italian citizens participated in all three assessments. The demographic composition of the sample is reported in Table 1. All the survey waves comprised an evaluation of sleep quality, insomnia symptoms, chronotype, depressive symptomatology, perceived stress, and anxiety through the following validated questionnaires: the Pittsburgh Sleep Quality Index (Curcio et al., 2013; PSQI), the Insomnia Severity Index (Castronovo et al., 2016; ISI), the Morningness-Eveningness Questionnaire-reduced version (Natale et al., 2006; MEQr), the Beck Depression Inventory-second edition (Sica and Ghisi, 2007; BDI-II), the 10-item Perceived Stress Scale (Mondo et al., 2021; PSS-10), and the state– anxiety subscale of the State-Trait Anxiety Inventory (Spielberger et al., 1970; STAI-X1). The administration order of mandatory sleep questionnaires was as follows: PSQI, ISI, and MEQr. Subsequently, participants could decide whether continue the compilation of the other three questionnaires (BDI-II, PSS-10, STAI-X1) with the option to stop after each of them to ensure reliable unforced responses due to the burden of testing battery. A total of 71, 43, and 40 subjects did non fill out the BDI-II in the first, second, and third survey wave, respectively. A total of 100, 61, and 83 participants did not complete the PSS-10 during the three assessment points, respectively. Finally, 103, 61, and 116 respondents did not fill out the STAI-X1 in the three survey waves. The Institutional Review Board of the University of L’Aquila approved the research project (protocol n. 43066/2020). The study was performed according to the principles established by the Declaration of Helsinki. Online informed consent was obtained from participants.

**Table 1.** Demographic characteristics of the sample participating in the three survey waves.

|  | April 2020<br>(survey wave 1) | December 2020<br>(survey wave 2) | April 2022<br>(survey wave 3) |
| --- | --- | --- | --- |
|  | N (%) or *mean (SD) |  |  |
| Age | *35.325 (12.59) | *35.985 (12.59) | *37.367 (12.59) |
| Gender |  |  |  |
| Male | 202 (19.02) | 202 (19.02) | 202 (19.02) |
| Female | 860 (80.98) | 860 (80.98) | 860 (80.98) |
| Education |  |  |  |
| Middle/High school | 318 (29.94) | 285 (26.84) | 238 (22.41) |
| Graduate | 599 (56.40) | 629 (59.23) | 633 (59.60) |
| Postgraduate | 145 (13.65) | 148 (13.94) | 191 (17.99) |
| Occupation |  |  |  |
| Unemployed | 81 (7.63) | 77 (7.25) | 95 (8.95) |
| Student | 305 (28.72) | 241 (22.69) | 167 (15.73) |
| Healthcare worker | 68 (6.40) | 90 (8.47) | 106 (9.98) |
| Self-employed | 198 (18.64) | 189 (17.80) | 212 (19.96) |
| Employed | 379 (35.69) | 428 (40.30) | 443 (41.71) |
| Retired | 31 (2.92) | 37 (3.48) | 39 (3.67) |

### Questionnaires

The PSQI (Curcio et al., 2013) is a widely used questionnaire to evaluate sleep quality. It consists of nineteen questions covering seven dimensions (subjective sleep quality, sleep duration, sleep latency, habitual sleep efficiency, sleep disturbances, the use of sleeping medications, and daytime dysfunctions). A higher score in each sub-component (range, 0–3) indicates greater severity of symptoms on the specific dimension (i.e., higher scores on *subjective sleep quality* sub-component corresponds to lower sleep quality, higher scores on *sleep duration* sub-component indicates shorter sleep duration, higher scores in *sleep latency* sub-component points to longer sleep latency, and so on), giving rise to a higher global score (range, 0–21) that suggests poorer sleep quality. The ISI (Castronovo et al., 2016) is a well-established 7-item tool to measure the severity of insomnia symptoms in clinical settings (range, 0–21). A higher score points to a more severe insomnia condition. The MEQr (Natale et al., 2006) is a 5-item instrument typically used in chrono-psychological research to estimate circadian typologies within the morningness-eveningness continuum. A lower total score (range, 4–25) suggests a tendency to eveningness and *vice versa* for morningness. The BDI-II (Sica & Ghisi, 2007) is a 21-item self-report inventory used in clinical practice to evaluate the severity of depression symptoms. A higher score (range, 0–63) is interpreted as a more severe depressive symptomatology. The PSS-10 (Mondo et al., 2021) is a 10-item questionnaire to assess thoughts and feelings about stressful events. A higher score (range, 0–40) indicates a higher level of perceived stress. The STAI-X1 (Spielberger et al., 1970) is a 20-item anxiety inventory belonging to the Cognitive Behavioural Assessment battery 2.0 (Sanavio et al., 1998). A higher total score points to more severe state anxiety.

### Statistical analysis

The statistical analyses were performed using the lme4 R package (Bates et al., 2015), which provides functions for fitting and analysing mixed models. Models were fitted using REML adopting the Satterthwaite approximation to compute *p*-values (Luke, 2017). Mixed-model analyses included a random intercept per participant to account for the repeated-measures nature of the data and the variability among respondents’ scores. Bonferroni post hoc tests and simple effect contrasts using the “emmeans” R package (Lenth et al., 2022) were computed in the case of significant main effects or interaction effects, respectively. The level of significance was always set at *p* < 0.05.

Firstly, we ran different models including the scores of each sleep and mental health questionnaire (PSQI, ISI, BDI-II, PSS-10, and STAI-X1) as dependent variables, and the *survey wave* factor (April 2020, December 2020, April 2022) as three-level within-subjects predictor. These analyses aimed to explore the general trajectories of sleep and psychological disturbances among the overall sample along the two pandemic years.

Furthermore, the same analysis was performed for specific items of PSQI [total sleep time (min), bedtime (hh:mm) and get-up time (hh:mm)] and each sub-component of PSQI (subjective sleep quality, sleep latency, sleep duration, habitual sleep efficiency, sleep disturbances, sleep medications, and daytime dysfunction) to describe the time course of specific dimensions of sleep habits/quality across the three assessments. Finally, we ran further mixed models on questionnaire scores (PSQI, ISI, BDI-II, PSS-10, and STAI-X1) including *survey wave* (April 2020, December 2020, April 2022), *age* (continuous time-varying variable), *gender* (male, female), *MEQr scores* (continuous time-varying variable), and the interaction of *survey wave* factor with *age, gender*, and *MEQr scores* as predictors. These analyses aimed at describing possible differences in the trajectories of sleep quality, insomnia, depression, perceived stress, and anxiety depending on the above-mentioned demographic and chrono-psychological factors. Continuous moderators (age and MEQr score) were represented by plotting mean ± standard deviation values to provide a graphical representation of significant interaction effects. As regards the age variable, we coded the mean age − 1 standard deviation (23.612 years) as “younger”, the mean age (36.226 years) as “middle-age”, and the mean age + 1 standard deviation (48.540 years) as “older”. As regards MEQr scores, we labelled the mean MEQr score − 1 standard deviation (11.803) as “evening-type”, the mean MEQr score (15.494) as “intermediate-type”, and the mean + 1 standard deviation (19.185) as “morning-type”. Due to the optional nature of BDI-II, PSS-10, and STAI-X1, Little’s MCAR test was performed using SPSS version 27.0.1.0 (IBM Corp, Armonk, NY, USA), which showed that missing data occurred completely at random over the three survey waves (all *p* > 0.241). Harman’s single factor test did not identify common method bias in each survey wave.

## Results

### Sleep variables

PSQI overall score did not differ between the three survey waves (F_2,2032.77_ = 1.637, *p* = 0.195), indicating that sleep quality was stable over the pandemic. On the other hand, the *Survey wave* factor was significant in the analyses on ISI score (F_2,2122_ = 28.699, *p* < 0.001) and total sleep time (F_2,2122_ = 47.751, *p* < 0.001). Bonferroni post hoc comparisons (Fig. 1) revealed a progressive improvement of insomnia symptomatology and decreased sleep duration. Specifically, the second and third survey waves were characterised by decreased ISI scores (−0.617, *p* < 0.001; −1.076, *p* < 0.001; respectively) and total sleep time (−15.395 min, *p* < 0.001; −20.706 min, *p* < 0.001; respectively) than the first one. The third assessment was associated with reduced ISI scores (−0.460, *p* = 0.004) and sleep duration (−5.311 min, *p* = 0.048) than the second one.

**Figure 1.**
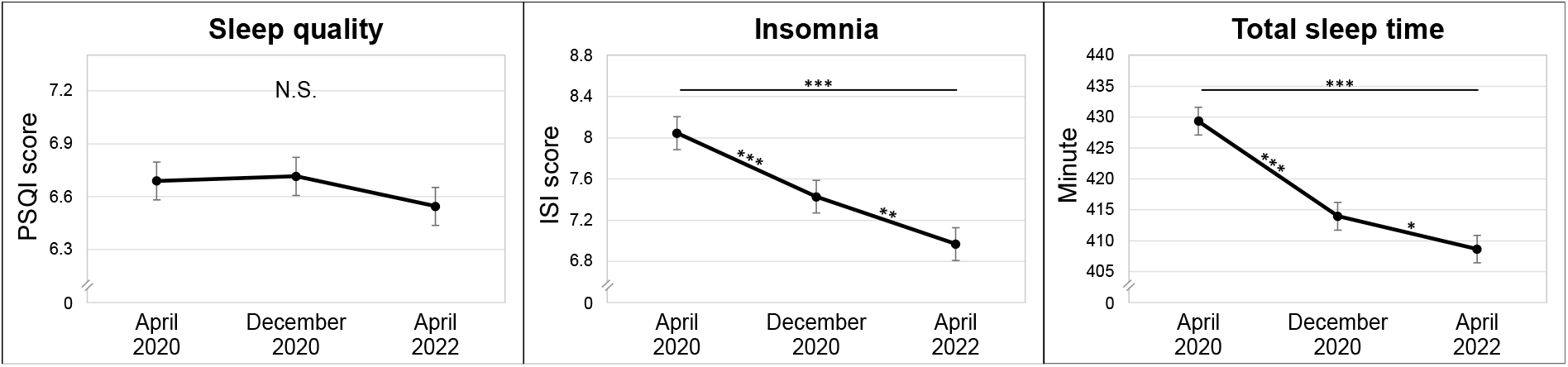
Mean ± standard error of PSQI scores (sleep quality), ISI scores (insomnia), and total sleep time (min) during the three survey waves. Significant Bonferroni post hoc comparisons are indicated with asterisks (* *p* < 0.05, ** *p* < 0.01, *** *p* < 0.001). *Abbreviations:* PSQI, Pittsburgh Sleep Quality Index; ISI, Insomnia Severity Index.

Analyses also showed a significant effect of the *survey wave* factor on bedtime (F_2,2122_ = 264.513, *p* < 0.001), and get-up time (F_2,2122_ = 430.555, *p* < 0.001). As shown in Fig. 2, post hoc comparisons indicated the progressive advance of bedtime and get-up time over the two pandemic years. Specifically, participants reported earlier bedtime and get-up time during the second (−34.567 min, *p* < 0.001; −50.400 min, *p* < 0.001; respectively) and third survey waves (−50.400 min, *p* < 0.001; −70.567 min, *p* < 0.001; respectively) than the first one. Bedtime and get-up time were also significantly advanced in the third assessment compared with the second one (−15.833 min, *p* < 0.001; −20.167 min, *p* < 0.001; respectively).

**Figure 2.**
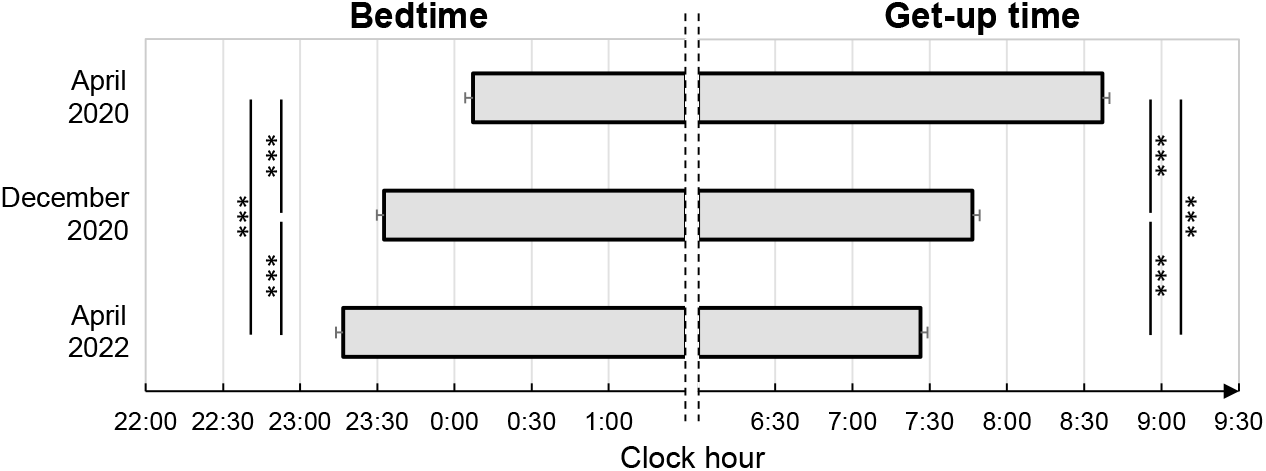
Mean ± standard error of bedtime and get-up time (hh:mm) during the three survey waves. Asterisks indicated significant Bonferroni post hoc comparisons (*** *p* < 0.001).

Results of the analyses on PSQI sub-components are reported in Table 2. We highlighted significant differences between survey waves on subjective sleep quality, sleep latency, sleep duration, sleep disturbances, and daytime dysfunction. Participants reported better subjective sleep quality and less severe sleep disturbances during the third assessment time than the first one (*p* < 0.001, *p* = 0.001, respectively) and the second one (*p* = 0.005, *p* = 0.046, respectively). Sleep onset latency was reduced over time (all *p* < 0.001). Conversely, post hoc comparisons showed reduced sleep duration during the second and the third survey waves compared to the first one (both *p* < 0.001) and increased daytime dysfunctions over time (all *p* < 0.028).

**Table 2.** Descriptive statistics [mean (SD)] of PSQI sub-component scores during the three survey waves (survey wave 1: April 2020; survey wave 2: December 2020; survey wave 3: April 2022) and the corresponding statistical comparisons (*F, p, Bonferroni post hoc*). A higher score in each sub-component indicates greater severity of the respective problem (see “Questionnaires” section for a detailed description of the sub-component score interpretation).

|  | April 2020 | December 2020 | April 2022 |  |  |  |
| --- | --- | --- | --- | --- | --- | --- |
| *PSQI sub-component* | ***Mean (SD)*** |  |  | ***F*** | ***p*** | ***Post hoc*** |
| Subjective sleep quality | 1.36 (0.76) | 1.30 (0.71) | 1.23 (0.66) | 14.72 | **<0.001** | SW1>SW3***<br>SW2>SW3** |
| Sleep latency | 1.36 (1.02) | 1.20 (1.01) | 1.07 (0.96) | 43.61 | **<0.001** | SW1>SW2>SW3*** |
| Sleep duration | 0.70 (0.79) | 0.81 (0.78) | 0.85 (0.77) | 21.01 | **<0.001** | SW1<SW2***<br>SW1<SW3*** |
| Habitual sleep efficiency | 0.77 (0.99) | 0.79 (0.98) | 0.80 (0.97) | 0.52 | 0.59 |  |
| Sleep disturbances | 1.39 (0.58) | 1.37 (0.56) | 1.32 (0.55) | 6.39 | **0.002** | SW1>SW3**<br>SW2>SW3* |
| Sleep medications | 0.26 (0.77) | 0.32 (0.85) | 0.29 (0.82) | 2.18 | 0.11 |  |
| Daytime dysfunction | 0.83 (0.71) | 0.90 (0.67) | 0.97 (0.71) | 15.00 | **<0.001** | SW1<SW2<SW3* |

Note: Significant values are in bold.
Abbreviations: SD, standard deviation; PSQI, Pittsburgh Sleep Quality Index; SW, Survey wave.

### Age, gender, and chronotype effect on sleep variables

As reported in Table 3, older age was associated with poorer sleep quality, more severe insomnia symptoms, and shorter total sleep time. The interaction between *survey wave* and *age* factors was significant in the analyses of all the sleep variables (PSQI score, ISI score, total sleep time), indicating an age-dependent time course of sleep quality, insomnia symptoms, and sleep duration.

**Table 3.**
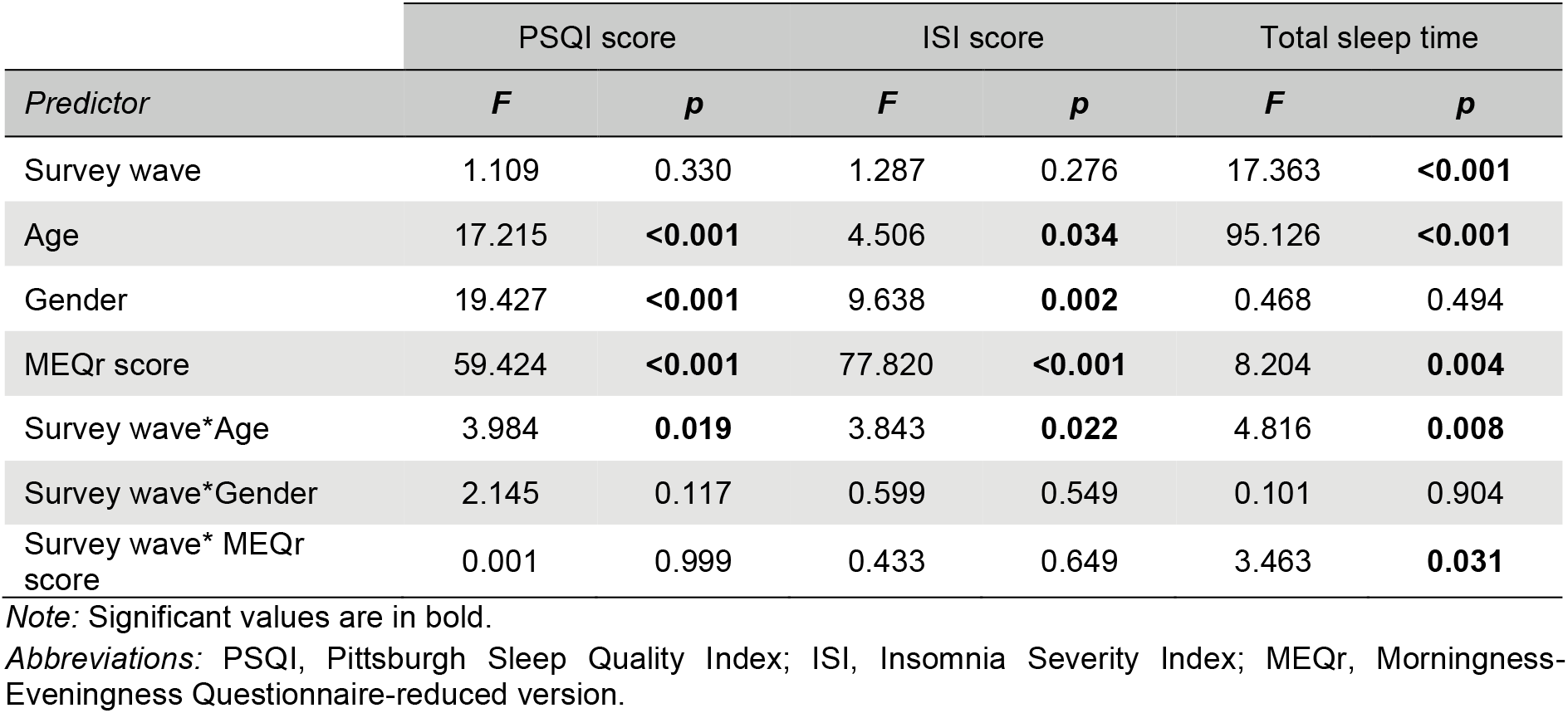
Results (*F* and *p*) of the mixed model analyses on PSQI score (sleep quality), ISI score (insomnia), and total sleep time (min). The models comprised the following predictors: survey wave (April 2020, December 2020, April 2022), age, gender (male, female), MEQr score (chronotype), and the interaction between survey wave and age, gender, and MEQr score.

| *Predictor* | PSQI score |  | ISI score |  | Total sleep time |  |
| --- | --- | --- | --- | --- | --- | --- |
|  | *F* | *p* | *F* | *p* | *F* | *p* |
| Survey wave | 1.109 | 0.330 | 1.287 | 0.276 | 17.363 | **<0.001** |
| Age | 17.215 | **<0.001** | 4.506 | **0.034** | 95.126 | **<0.001** |
| Gender | 19.427 | **<0.001** | 9.638 | **0.002** | 0.468 | 0.494 |
| MEQr score | 59.424 | **<0.001** | 77.820 | **<0.001** | 8.204 | **0.004** |
| Survey wave*Age | 3.984 | **0.019** | 3.843 | **0.022** | 4.816 | **0.008** |
| Survey wave*Gender | 2.145 | 0.117 | 0.599 | 0.549 | 0.101 | 0.904 |
| Survey wave* MEQr score | 0.001 | 0.999 | 0.433 | 0.649 | 3.463 | **0.031** |

*Note:* Significant values are in bold.
*Abbreviations:* PSQI, Pittsburgh Sleep Quality Index; ISI, Insomnia Severity Index; MEQr, Morningness-Eveningness Questionnaire-reduced version.

Simple effect contrasts (Fig. 3) highlighted significant differences between the survey waves among the older respondents, as shown by increased PSQI scores in December 2020 (+0.336, *p* = 0.041), and a subsequent significant improvement in sleep quality during the last assessment period (−0.369, *p* = 0.020). No differences were reported by the middle-age (+0.096, *p* = 0.452) and the younger participants (−0.144, *p* = 0.385) during the second survey compared to the first one, and from the second to the third assessment (−0.104, *p* = 0.417; +0.162, *p* = 0.338; respectively).

**Figure 3.**
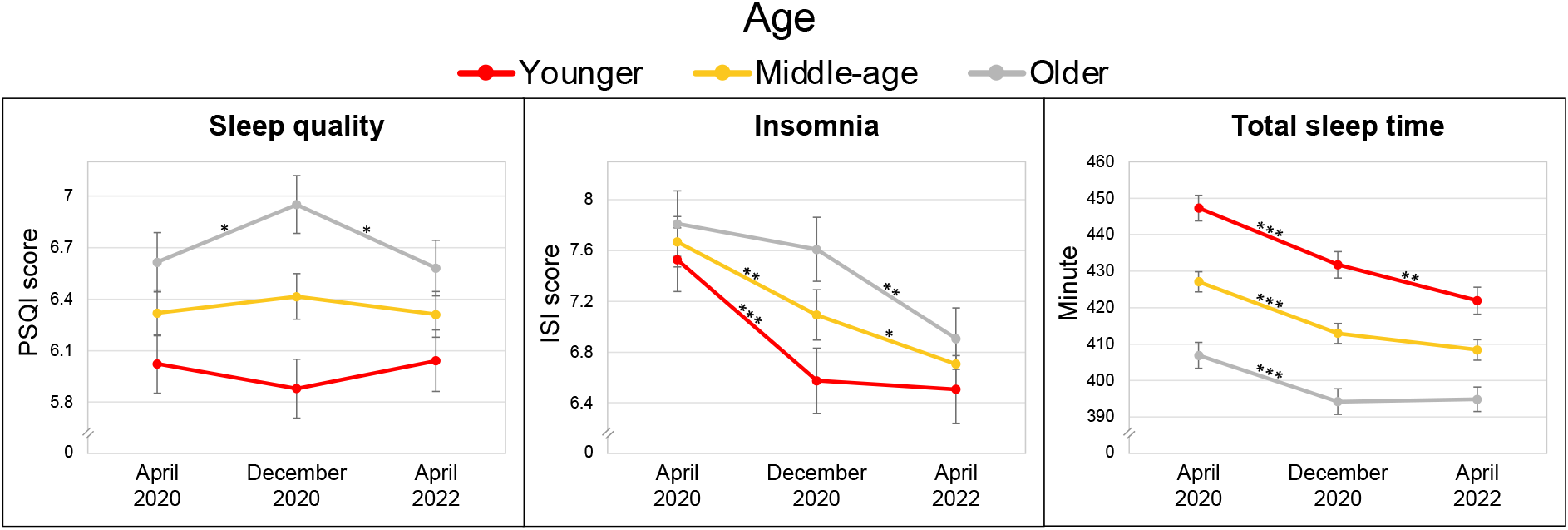
Estimated marginal mean ± standard error of PSQI scores (sleep quality), ISI score (insomnia), and total sleep time (min) during the three survey waves according with age (red: young, yellow: middle-age, grey: older). Significant simple effect contrasts are indicated with asterisks (* *p* < 0.05, ** *p* < 0.01, *** *p* < 0.001). *Abbreviations:* PSQI, Pittsburgh Sleep Quality Index; ISI, Insomnia Severity Index.

ISI scores decreased during the second survey wave compared to the first one in the middle-age (−0.577, *p* = 0.001) and young people (−0.952, *p* < 0.001), while insomnia was stable among the older respondents (−0.202, *p* = 0.384). On the other hand, older participants reported a significant reduction in ISI scores from the second to the third survey wave (−0.703, *p* = 0.002). A further decline in ISI scores was observed in April 2022 in the middle-age group compared to December 2020 (−0.386, *p* = 0.033), while the younger population did not show any change in insomnia symptoms comparing the last two survey waves (−0.068, *p* = 0.776). Finally, an overall reduction in total sleep time was observed in December 2020 compared to April 2020 (young: −15.551 min; middle-age: −14.122 min; older: −12.694 min; all *p* < 0.001), and the young population was marked by a further reduction in sleep time during the last assessment (−9.801 min, *p* = 0.008). Older and middle-age respondents reported stable sleep duration in the last two survey waves (+0.635 min, *p* = 0.855; −4.583 min, *p* = 0.103)

Female participants reported poorer sleep quality and more severe insomnia symptoms than males (Table 3). No differences in total sleep time between genders emerged. The interaction between *survey wave* and *gender* factors on sleep variables did not reveal differences between genders in the time course of sleep quality, insomnia symptoms, and sleep duration. Finally, eveningness was associated with lower sleep quality, more severe insomnia symptomatology, and shorter total sleep time (Table 3). The interaction between *survey wave* and *MEQr scores* in predicting sleep duration was significant, while no different time course of sleep quality and insomnia symptoms between circadian typologies was highlighted. As shown in Fig. 4, simple effect contrasts revealed a larger reduction of total sleep time among the evening-type population (−17.068 min, *p* < 0.001) from the first to the second survey wave than among the intermediate-type (−14.122, *p* < 0.001) and morning-type respondents (−11.177, *p* = 0.002). Late chronotypes are associated with a further significant reduction of sleep time from the second to the third assessment point (−7.774 min, *p* = 0.028), while no significant differences were reported by intermediate-type (−4.583 min, *p* = 0.102) and morning-type participants (−1.392 min, *p* = 0.708).

**Figure 4.**
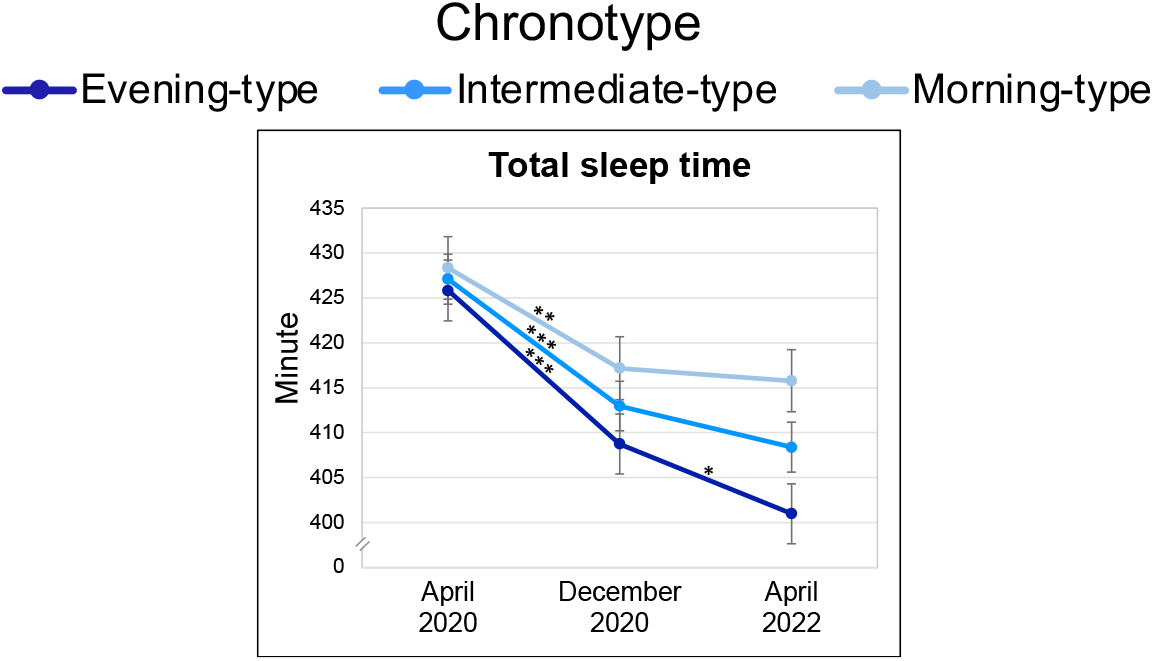
Estimated marginal mean ± standard error of total sleep time (min) during the three survey waves according with chronotype (light blue: morning-type, blue: intermediate-type, dark blue: evening-type). Significant simple effect contrasts are indicated with asterisks (* *p* < 0.05, *** *p* < 0.001).

### Psychological variables

The analyses on BDI-II, PSS-10, and STAI-X1 scores showed significant differences between the three survey waves (F_2,2016.04_ = 4.142, *p* = 0.016; F_2,1974.17_ = 16.576, *p* < 0.001; F_2,1911.25_ = 43.850, *p* < 0.001, respectively). As shown in Fig. 5, participants reported less severe depressive symptoms during the third survey wave than the first one (−0.661, *p* = 0.023), while the second assessment did not significantly differ from the first (−0.109, *p* = 1.000) and the third ones (+0.552, *p* = 0.074). Perceived stress increased during the second (+1.043, *p* < 0.001) and third survey waves (+1.011, *p* < 0.001) than the first one. Anxiety scores began to decline in December 2020 (−1.887, *p* < 0.001) and were further reduced during the last assessment compared to the second one (−0.904, *p* = 0.008).

**Figure 5.**
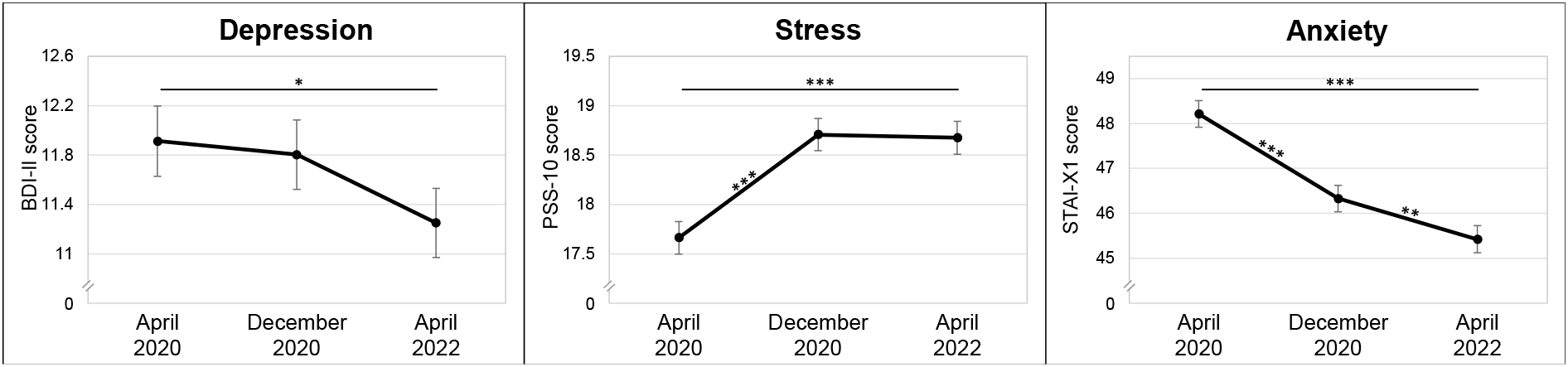
Mean ± standard error of BDI-II scores (depression), PSS-10 scores (perceived stress), and STAI-X1 scores (anxiety), during the three survey waves. Significant Bonferroni post hoc comparisons are indicated with asterisks (* *p* < 0.05, ** *p* < 0.01, *** *p* < 0.001). *Abbreviations:* BDI-II, Beck Depression Inventory-second edition; PSS-10, Perceived Stress Scale-10 item; STAI-X1, state-anxiety subscale of the State-Trait Anxiety Inventory.

### Age, gender, and chronotype effect on psychological variables

Younger age, female gender, and eveningness were associated with more severe depressive symptoms and higher perceived stress. Younger respondents and evening-type people reported higher anxiety levels, while no relationship between age and anxiety emerged (Table 4). The interaction between *survey wave* and chrono-demographic factors (*age, gender, MEQr score*) in predicting PSS-10 was significant. No significant interaction effect for BDI-II and STAI-X1 scores was highlighted, indicating no differences between age groups, genders, and chronotypes in the time course of depressive and anxiety symptoms.

**Table 4.** Results (*F* and *p*) of the mixed model analyses on BDI-II score (depression), PSS-10 score (perceived stress), and STAI-X1 score (anxiety). The models comprised the following predictors: survey wave (April 2020, December 2020, April 2022), age, gender (male, female), MEQr score (chronotype), and the interaction between survey wave and age, gender, and MEQr score.

| *Predictor* | BDI-II score |  | PSS-10 score |  | STAI-X1 score |  |
| --- | --- | --- | --- | --- | --- | --- |
|  | *F* | *p* | *F* | *p* | *F* | *p* |
| Survey wave | 0.949 | 0.387 | 26.924 | **<0.001** | 27.373 | **<0.001** |
| Age | 8.137 | **0.004** | 15.065 | **<0.001** | 0.134 | 0.714 |
| Gender | 17.034 | **<0.001** | 30.345 | **<0.001** | 14.842 | **<0.001** |
| MEQr score | 81.224 | **<0.001** | 30.133 | **<0.001** | 25.545 | **<0.001** |
| Survey wave*Age | 0.404 | 0.668 | 7.072 | **<0.001** | 0.061 | 0.941 |
| Survey wave*Gender | 1.330 | 0.265 | 12.596 | **<0.001** | 0.644 | 0.525 |
| Survey wave* MEQr score | 0.484 | 0.616 | 3.646 | **0.026** | 1.182 | 0.307 |

*Note:* Significant values are in bold.
*Abbreviations:* BDI-II, Beck Depression Inventory-second edition; PSS-10, Perceived Stress Scale-10 item; STAI-X1, state-anxiety subscale of the State-Trait Anxiety Inventory; MEQr, Morningness-Eveningness Questionnaire-reduced version.

Simple effect contrasts (Fig. 6) revealed significantly raised stress levels from the first survey wave to the second one in all groups. However, the extent of the effect depended on age, gender, and chronotype. Specifically, older participants and morning-type subjects reported the largest increase in perceived stress (+2.461, *p* < 0.001; +2.145, *p* < 0.001; respectively), middle-age and intermediate-type respondents showed an intermediate increase (+1.657, *p* < 0.001; +1.657, *p* < 0.001; respectively), while younger and evening-type subjects were associated with the smallest increase (+0.853, *p* = 0.010; +1.169, *p* < 0.001; respectively). Men reported a greater increase in perceived stress (+2.497, *p* < 0.001) than women (+0.817, *p* < 0.001). Finally, all groups reported unchanged stress levels between the second and the third survey waves (younger: −0.205, *p* = 0.546; middle-age: −0.012, *p* = 0.962; older: +0.180, *p* = 0.572; male: +0.001, *p* = 0.999; female: −0.025, *p* = 0.911; evening-type: −0.038, *p* = 0.907; intermediate-type: −0.012, *p* = 0.962; morning-type: +0.013, *p* = 0.969).

**Figure 6.**
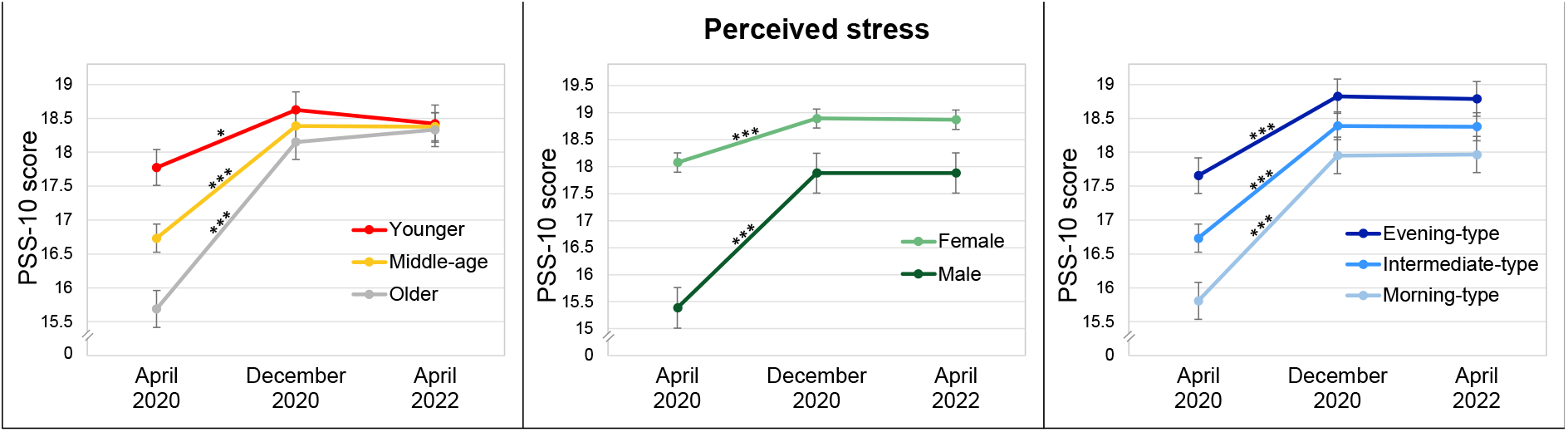
Estimated marginal mean ± standard error of PSS-10 scores (perceived stress) during the three survey waves according with age (red: young, yellow: middle-age, grey: older), gender (light green: female, dark green: male), and chronotype (light blue: morning-type, blue: intermediate-type, dark blue: evening-type). Significant simple effect contrasts are indicated with asterisks (* *p* < 0.05, *** *p* < 0.001). *Abbreviations:* PSS-10, Perceived Stress Scale-10 item.

## Discussion

The present study demonstrated different trajectories of sleep quality, insomnia, depression, stress, and anxiety during the two pandemic years.

As indicated by the PSQI total score, the overall sleep quality was stable across the three assessments. However, an overview of PSQI sub-components suggested a more articulated pattern of results, revealing a clear dissociation of the effects on sleep features depending on the specific domain considered. On the one hand, subjective sleep quality improved, and sleep disturbances and sleep onset latency decreased two years after the lockdown of Spring 2020. These findings are consistent with the progressive reduction of insomnia symptoms reported by the overall sample. Conversely, we demonstrated a gradual decrease in sleep duration and an increase in sleep-related daytime dysfunctions with the loosening of the restraining measures. Similar results were obtained by our previous investigation (Salfi, D’Atri, et al., 2021) comparing the lockdown scenario and the second contagion wave of COVID-19 and by other studies carried out in the post-lockdown period (Alfonsi et al., 2021; Massar et al., 2021; Salfi, D’Atri, et al., 2021). These results could be ascribable to the peculiar nature of the lockdown. The stressful situation led to worsened sleep disturbances worldwide (Jahrami et al., 2021, 2022). Meanwhile, the strict confinement regimen was associated with unlocked time for sleep and reduced social jetlag for most of the population (Kantermann, 2020; Korman et al., 2020; Leone et al., 2020), with a consequent improvement in sleep-related daytime dysfunctions. However, social and working obligations returned to exert their negative effect on sleep during the second survey wave, and even more two years after the lockdown. Consistently, participants reported substantial earlier get-up times (∼70 min) and a smaller but still important advanced bedtime (∼50 min) in April 2022 compared to the first lockdown. The discrepancy between the above-mentioned variations led to an overall reduction in total sleep time, as earlier awakenings were not adequately compensated by a comparable advance in bedtimes.

The improvement in sleep disturbances was accompanied by reduced depressive and anxiety symptoms. However, as far as depression is concerned, we observed a marginal decrease in the BDI-II score, whose effect reached significance only comparing the first and the third assessment. The time course of self-perceived stress was in stark contrast to the other examined psychological dimensions. The stress level increased from the lockdown period, reaching its plateau during the second contagion wave, and then stabilised. As suggested by other investigations (Conte et al., 2021; Salfi, D’Atri, et al., 2021), this outcome could be ascribable to the prolonged nature of the emergency, which was accompanied by a societal and economic crisis. Moreover, after lifting restrictions, the infection risk raised, presumably affecting the perceived stress in the general population.

Demographic factors such as age also influenced the observed effects. Older participants reported poorer sleep quality, more severe insomnia symptoms, and shorter sleep duration than younger respondents. This outcome is consistent with the pre-pandemic (Kamel & Gammack, 2006; Madrid-Valero et al., 2017) literature and is related to the typical sleep changes occurring across the lifespan (Li et al., 2018; Patel et al., 2018). Moreover, in line with studies performed during the current period (Amicucci et al., 2021; Manchia et al., 2022; Rossi et al., 2020), the younger population showed more severe depressive symptoms and no difference in anxiety. Similar trajectories of depression and anxiety were observed over the three assessment points among the different age groups. However, our investigation highlighted an age-dependent time course of sleep quality, insomnia, total sleep time, and perceived stress. Younger and middle-age groups reported unchanged overall sleep quality over the two pandemic years. On the other hand, we observed reduced sleep quality during the second assessment in older participants and a subsequent improvement during the third survey wave. Moreover, the three age groups seemed to report a similar extent of insomnia relief two years after the first lockdown, but this effect was reached at different time points. Younger people reported the maximum improvement already during Winter 2020, middle-age participants gradually improved across the three survey waves, while insomnia symptoms of older respondents were stable until December 2020 and then improved. Finally, we confirmed higher stress levels among young people during the lockdown (Amicucci et al., 2021; Rossi et al., 2020; Ueda et al., 2020), while we reported a general increase in perceived stress among all age groups, which led to similar stress levels in the overall sample during the second and third survey waves.

The specific characteristics of the pandemic scenario during the three data collections could explain our pattern of results. During the second contagion wave of Winter 2020, the confinement measures were reduced, several business and school activities reopened, and freedom of movement was partially ensured. Therefore, infection risk increased while no vaccine against COVID-19 was still available. Considering that older age is associated with the highest morbidity and mortality rates due to SARS-CoV-2 infection, it is unsurprising that older respondents experienced a substantial increase in stress levels with a concurrent decline in sleep quality and unchanged insomnia severity in December 2020 compared to the first lockdown period. However, the massive vaccination campaign of the subsequent months and the return to an almost normal life after two years could have allowed older participants to experience improved sleep disturbances. On the other hand, several studies suggested that the younger population suffered particularly from the collapse of social interactions during the confinement regimens (Elmer et al., 2020; Sampogna et al., 2021; Viselli et al., 2021; Weissbourd et al., 2020) with well-documented repercussions on sleep and mental health (Amicucci et al., 2021; Daly et al., 2020; Jahrami et al., 2022; Rossi et al., 2020). Consequently, the loosening of confinement measures and the partial resumption of social interactions could explain the small increase in stress and the concomitant improvement of insomnia symptomatology among the younger people already during Winter 2020.

The present investigation confirmed the vulnerability of women to experience sleep ad psychological problems during the pandemic (Daly et al., 2020; Rossi et al., 2020; Salfi et al., 2020; Salfi, Lauriola, et al., 2021). However, we did not detect differences between genders in the trajectories of sleep quality, insomnia, depression, and anxiety symptoms across the three survey waves. Furthermore, our investigation showed that men reported definitely less stress than women during the first weeks of lockdown, although this difference was largely reduced since Winter 2020. This finding is consistent with the interpretation that females might have already reached the peak of psychological distress during the first weeks of home confinement (Salfi et al., 2020). Conversely, it seems that men reacted better to the lockdown, but the prolonged emergency led them to experience higher distress in the long run.

Finally, our study confirmed the well-known vulnerability to sleep disturbances and psychological problems of evening-type people (Adan et al., 2012). This evidence was also reported by investigations carried out during the first stage of the pandemic (Merikanto et al., 2022; Salfi, Lauriola, et al., 2021). All chronotypes were associated with a similar time course of sleep quality, insomnia, depression, stress, and anxiety across the three periods covered by our study. Meanwhile, the changes in sleep duration differed between circadian typologies. During the lockdown, the typical discrepancy in sleep duration between chronotypes disappeared. However, the progression of the pandemic and the gradual resumptions of social and working obligations reinstated the well-documented misalignment between the evening-oriented biological clock and the morning-oriented social schedule of the so-called “night-owls” (Roenneberg et al., 2019). Consequently, this situation led the evening-type participants to sleep less and less as the COVID-19 crisis improved and people resumed their pre-existing daily routine (Salfi et al., 2022).

## Conclusions

Since the first months of 2020, the COVID-19 has pervasively affected every area of life of the worldwide population. A massive amount of literature has been developed during the lockdown period, confirming pervasive repercussions on the sleep and mental health of the general population. The second contagion wave of COVID-19 continued to be characterised by psychological distress and impaired sleep health worldwide.

To the best of our knowledge, the present investigation is the first to longitudinally examine the long-term trajectories of sleep quality/habits, insomnia, depression, stress, and anxiety after two years from the lockdown, providing novel insights on the time course of sleep and mental health according with age, gender, and circadian typology across three critical stages of the COVID-19 outbreak. However, some limitations should be acknowledged. We analysed a large sample of Italian participants. Nevertheless, the sample comprised a higher prevalence of women, and participants were recruited by adopting a non-probabilistic sampling technique. Finally, the evaluation of sleep features, chronotype, and psychological well-being relied on self-reported questionnaires. In this view, caution is required in generalising the present findings due to possible selection and response biases which could affect our data.

In conclusion, our study described a promising scenario after two years of the pandemic. We demonstrated decreased sleep disturbances, insomnia, depressive, and anxiety symptoms. However, the extent of the improvements was relatively small. Meanwhile, the re-establishment of pre-pandemic social and working dynamics configured a negative effect on sleep duration, which was reduced among the overall sample, and more strongly in particular population groups such as younger and evening-type people. Finally, the persistence of high stress levels and the decreased distress differences between age groups, genders, and chronotypes suggest that people transversely continued to feel the burden of this unprecedented and protracted historical period. In this view, further long-term monitoring of sleep and mental health time course is necessary to claim the end of the COVID-19 emergency on sleep and psychological status of the general population.

## Data Availability

All data produced in the present study are available upon reasonable request to the authors

## Data availability statement

The data supporting the findings of this study are available from the corresponding author upon reasonable request.

## Notes

**Conflict of interests:** None of the authors have potential conflicts of interest to be disclosed. All authors have seen and approved the manuscript.

### Competing Interest Statement

The authors have declared no competing interest.

### Funding Statement

This study did not receive any funding

### Author Declarations

IRB of University of L Aquila gave ethical approval for this work (protocol n. 43066).

## References

Adan, A., Archer, S. N., Hidalgo, M. P., di Milia, L., Natale, V., & Randler, C. (2012). Circadian typology: A comprehensive review. Chronobiology International, 29(9), 1153–1175. https://doi.org/10.3109/07420528.2012.719971

Alfonsi, V., Gorgoni, M., Scarpelli, S., Zivi, P., Sdoia, S., Mari, E., Fraschetti, A., Ferlazzo, F., Giannini, A. M., & de Gennaro, L. (2021). COVID-19 lockdown and poor sleep quality: Not the whole story. Journal of Sleep Research, 30(5), 1–6. https://doi.org/10.1111/jsr.13368

Amicucci, G., Salfi, F., D’atri, A., Viselli, L., & Ferrara, M. (2021). The Differential Impact of COVID-19 Lockdown on Sleep Quality, Insomnia, Depression, Stress, and Anxiety among Late Adolescents and Elderly in Italy. Brain Sciences, 11(10). https://doi.org/10.3390/BRAINSCI11101336

Basishvili, T., Oniani, N., Sakhelashvili, I., Eliozishvili, M., Khizanashvili, M., Arabidze, M., Tsaava, M., Charekishvili, T., Tsertsvadze, N., & Darchia, N. (2021). Insomnia, Pre-Sleep Arousal, Psychosocial Factors and Changes in Sleep Pattern during the Second Wave Lockdown of the COVID-19 Pandemic in Georgia. Brain Sciences, 12(1), 17. https://doi.org/10.3390/brainsci12010017

Bates, D., Mächler, M., Bolker, B. M., & Walker, S. C. (2015). Fitting Linear Mixed-Effects Models Using lme4. Journal of Statistical Software, 67(1), 1–48. https://doi.org/10.18637/JSS.V067.I01

Benke, C., Autenrieth, L. K., Asselmann, E., & Pané-Farré, C. A. (2022). One year after the COVID-19 outbreak in Germany: long-term changes in depression, anxiety, loneliness, distress and life satisfaction. European Archives of Psychiatry and Clinical Neuroscience, 0123456789, 20–23. https://doi.org/10.1007/s00406-022-01400-0

Bottary, R., Fields, E. C., Kensinger, E. A., & Cunningham, T. J. (2022). Age and chronotype influenced sleep timing changes during the first wave of the COVID-19 pandemic. Journal of Sleep Research, 31(2), 1–7. https://doi.org/10.1111/jsr.13495

Casagrande, M., Favieri, F., Tambelli, R., & Forte, G. (2020). The enemy who sealed the world: effects quarantine due to the COVID-19 on sleep quality, anxiety, and psychological distress in the Italian population. Sleep Medicine, 75, 12–20. https://doi.org/10.1016/J.SLEEP.2020.05.011

Castronovo, V., Galbiati, A., Marelli, S., Brombin, C., Cugnata, F., Giarolli, L., Anelli, M. M., Rinaldi, F., & Ferini-Strambi, L. (2016). Validation study of the Italian version of the Insomnia Severity Index (ISI). Neurological Sciences, 37(9), 1517–1524. https://doi.org/10.1007/s10072-016-2620-z

Cellini, N., Canale, N., Mioni, G., & Costa, S. (2020). Changes in sleep pattern, sense of time and digital media use during COVID-19 lockdown in Italy. Journal of Sleep Research, 29(4), 1–6. https://doi.org/10.1111/jsr.13074

Chodkiewicz, J., Miniszewska, J., Krajewska, E., & Biliński, P. (2021). Mental health during the second wave of the covid-19 pandemic—polish studies. International Journal of Environmental Research and Public Health, 18(7). https://doi.org/10.3390/ijerph18073423

Conte, F., Cellini, N., de Rosa, O., Rescott, M. L., Malloggi, S., Giganti, F., & Ficca, G. (2021). Dissociated profiles of sleep timing and sleep quality changes across the first and second wave of the COVID-19 pandemic. Journal of Psychiatric Research, 143(September), 222–229. https://doi.org/10.1016/j.jpsychires.2021.09.025

Curcio, G., Tempesta, D., Scarlata, S., Marzano, C., Moroni, F., Rossini, P. M., Ferrara, M., & de Gennaro, L. (2013). Validity of the Italian Version of the Pittsburgh Sleep Quality Index (PSQI). Neurological Sciences, 34(4), 511–519. https://doi.org/10.1007/s10072-012-1085-y

Daly, M., & Robinson, E. (2022). Psychological distress associated with the second COVID-19 wave: Prospective evidence from the UK Household Longitudinal Study. Journal of Affective Disorders, 310(April), 274–278. https://doi.org/10.1016/j.jad.2022.05.025

Daly, M., Sutin, A., & Robinson, E. (2020). Longitudinal changes in mental health and the COVID-19 pandemic: Evidence from the UK Household Longitudinal Study. Psychological Medicine. https://doi.org/10.1017/S0033291720004432

Elmer, T., Mepham, K., & Stadtfeld, C. (2020). Students under lockdown: Comparisons of students’ social networks and mental health before and during the COVID-19 crisis in Switzerland. PLOS ONE, 15(7), e0236337. https://doi.org/10.1371/JOURNAL.PONE.0236337

Jahrami, H., Alhaj, O. A., Humood, A. M., Alenezi, A. F., Fekih-Romdhane, F., AlRasheed, M. M., Saif, Z. Q., Bragazzi, N. L., Pandi-Perumal, S. R., BaHammam, A. S., & Vitiello, M. v. (2022). Sleep disturbances during the COVID-19 pandemic: A systematic review, meta-analysis, and meta-regression. Sleep Medicine Reviews, 62, 101591. https://doi.org/10.1016/j.smrv.2022.101591

Jahrami, H., BaHammam, A. S., Bragazzi, N. L., Saif, Z., Faris, M., & Vitiello, M. v. (2021). Sleep problems during the COVID-19 pandemic by population: A systematic review and meta-analysis. Journal of Clinical Sleep Medicine, 17(2), 299–313. https://doi.org/10.5664/JCSM.8930

Kamel, N. S., & Gammack, J. K. (2006). Insomnia in the elderly: cause, approach, and treatment. The American Journal of Medicine, 119(6), 463–469. https://doi.org/10.1016/J.AMJMED.2005.10.051

Kantermann, T. (2020). Behavior: How a Global Social Lockdown Unlocks Time for Sleep. Current Biology, 30(14), R822–R823. https://doi.org/10.1016/j.cub.2020.06.037

Korman, M., Tkachev, V., Reis, C., Komada, Y., Kitamura, S., Gubin, D., Kumar, V., & Roenneberg, T. (2020). COVID-19-mandated social restrictions unveil the impact of social time pressure on sleep and body clock. Scientific Reports, 10(1), 1–10. https://doi.org/10.1038/s41598-020-79299-7

Lenth, R. v, Buerkner, P., Herve, M., Love, J., Miguez, F., Riebl, H., & Singmann, H. (2022). Package ‘emmeans’: Estimated Marginal Means, aka Least-Squares Means. Version 1.7.5. https://cran.r-project.org/web/packages/emmeans/emmeans.pdf

Leone, M. J., Sigman, M., & Golombek, D. A. (2020). Effects of lockdown on human sleep and chronotype during the COVID-19 pandemic. Current Biology, 30(16), R930–R931. https://doi.org/10.1016/j.cub.2020.07.015

Li, J., Vitiello, M. v., & Gooneratne, N. S. (2018). Sleep in Normal Aging. Sleep Medicine Clinics, 13(1), 1. https://doi.org/10.1016/J.JSMC.2017.09.001

Liu, Y., Wang, X., Sun, P., Zhang, Q., Zhang, C., Shen, Y., Wang, S., Ma, J., & Wang, G. (2022). Sleep disturbance and anxiety symptom among public during the second wave of COVID-19 in Beijing: A web-based cross-sectional survey. Journal of Affective Disorders, 298(PA), 80–85. https://doi.org/10.1016/j.jad.2021.10.068

Luke, S. G. (2017). Evaluating significance in linear mixed-effects models in R. Behavior Research Methods, 49(4), 1494–1502. https://doi.org/10.3758/S13428-016-0809-Y

Madrid-Valero, J. J., Martínez-Selva, J. M., Ribeiro do Couto, B., Sánchez-Romera, J. F., & Ordoñana, J. R. (2017). Age and gender effects on the prevalence of poor sleep quality in the adult population. Gaceta Sanitaria, 31(1), 18–22. https://doi.org/10.1016/J.GACETA.2016.05.013

Manchia, M., Gathier, A. W., Yapici-Eser, H., Schmidt, M. v., de Quervain, D., van Amelsvoort, T., Bisson, J. I., Cryan, J. F., Howes, O. D., Pinto, L., van der Wee, N. J., Domschke, K., Branchi, I., & Vinkers, C. H. (2022). The impact of the prolonged COVID-19 pandemic on stress resilience and mental health: A critical review across waves. European Neuropsychopharmacology, 55, 22–83. https://doi.org/10.1016/J.EURONEURO.2021.10.864

Massar, S. A. A., Ng, A. S. C., Soon, C. S., Ong, J. L., Chua, X. Y., Chee, N. I. Y. N., Lee, T. S., & Chee, M. W. L. (2021). Reopening after lockdown: The influence of working-from-home and digital device use on sleep, physical activity, and wellbeing following COVID-19 lockdown and reopening. Sleep. https://doi.org/10.1093/SLEEP/ZSAB250

Merikanto, I., Kortesoja, L., Benedict, C., Chung, F., Cedernaes, J., Espie, C. A., Morin, C. M., Dauvilliers, Y., Partinen, M., de Gennaro, L., Wing, Y. K., Chan, N. Y., Inoue, Y., Matsui, K., Holzinger, B., Plazzi, G., Mota-Rolim, S. A., Leger, D., Penzel, T., & Bjorvatn, B. (2022). Evening-types show highest increase of sleep and mental health problems during the COVID-19 pandemic - Multinational study on 19 267 adults. Sleep, 45(2), 1–13. https://doi.org/10.1093/sleep/zsab216

Mondo, M., Sechi, C., & Cabras, C. (2021). Psychometric evaluation of three versions of the Italian Perceived Stress Scale. Current Psychology, 40(4), 1884–1892. https://doi.org/10.1007/S12144-019-0132-8/TABLES/5

Natale, V., Esposito, M. J., Martoni, M., & Fabbri, M. (2006). Validity of the reduced version of the Morningness-Eveningness Questionnaire. Sleep and Biological Rhythms, 4(1), 72–74. https://doi.org/10.1111/j.1479-8425.2006.00192.x

Patel, D., Steinberg, J., & Patel, P. (2018). Insomnia in the Elderly: A Review. Journal of Clinical Sleep Medicine : JCSM : Official Publication of the American Academy of Sleep Medicine, 14(6), 1017–1024. https://doi.org/10.5664/JCSM.7172

Robinson, E., Sutin, A. R., Daly, M., & Jones, A. (2022). A systematic review and meta-analysis of longitudinal cohort studies comparing mental health before versus during the COVID-19 pandemic in 2020. Journal of Affective Disorders, 296(September 2021), 567–576. https://doi.org/10.1016/j.jad.2021.09.098

Roenneberg, T., Pilz, L. K., Zerbini, G., & Winnebeck, E. C. (2019). Chronotype and social jetlag: A (self-) critical review. Biology, 8(3), 1–19. https://doi.org/10.3390/biology8030054

Rossi, R., Socci, V., Talevi, D., Mensi, S., Niolu, C., Pacitti, F., di Marco, A., Rossi, A., Siracusano, A., & di Lorenzo, G. (2020). COVID-19 Pandemic and Lockdown Measures Impact on Mental Health Among the General Population in Italy. Frontiers in Psychiatry, 11, 790. https://doi.org/10.3389/FPSYT.2020.00790/BIBTEX

Rus Prelog, P., Matić, T., Pregelj, P., & Sadikov, A. (2022). Risk of Depression, Anxiety, and Stress During the Second Wave of COVID-19 in Slovenia. Frontiers in Psychiatry, 12(January), 1–9. https://doi.org/10.3389/fpsyt.2021.788898

Salfi, F., D’Atri, A., Amicucci, G., Viselli, L., Gorgoni, M., Scarpelli, S., Alfonsi, V., & Ferrara, M. (2022). The fall of vulnerability to sleep disturbances in evening chronotypes when working from home and its implications for depression. Scientific Reports, 12, 12249. https://doi.org/10.1038/s41598-022-16256-6

Salfi, F., D’Atri, A., Tempesta, D., & Ferrara, M. (2021). Sleeping under the waves: A longitudinal study across the contagion peaks of the COVID-19 pandemic in Italy. Journal of Sleep Research, 30(5). https://doi.org/10.1111/jsr.13313

Salfi, F., Lauriola, M., Amicucci, G., Corigliano, D., Viselli, L., Tempesta, D., & Ferrara, M. (2020). Gender-related time course of sleep disturbances and psychological symptoms during the COVID-19 lockdown: A longitudinal study on the Italian population. Neurobiology of Stress, 13, 100259. https://doi.org/10.1016/J.YNSTR.2020.100259

Salfi, F., Lauriola, M., D’Atri, A., Amicucci, G., Viselli, L., Tempesta, D., & Ferrara, M. (2021). Demographic, psychological, chronobiological, and work-related predictors of sleep disturbances during the COVID-19 lockdown in Italy. Scientific Reports, 11(1), 1–12. https://doi.org/10.1038/s41598-021-90993-y

Sampogna, G., Giallonardo, V., del Vecchio, V., Luciano, M., Albert, U., Carmassi, C., Carrà, G., Cirulli, F., Dell’Osso, B., Menculini, G., Belvederi Murri, M., Pompili, M., Sani, G., Volpe, U., Bianchini, V., & Fiorillo, A. (2021). Loneliness in Young Adults During the First Wave of COVID-19 Lockdown: Results From the Multicentric COMET Study. Frontiers in Psychiatry, 12, 2226. https://doi.org/10.3389/FPSYT.2021.788139/BIBTEX

Sanavio, E., Bertolotti, G., Michelin, P., Vidotto, G., & Zotti, A. M. (1998). CBA 2.0 – Cognitive Behavioural Assessment 2.0 - Scale Primarie: Manuale. Organizzazioni Speciali.

Sica, C., & Ghisi, M. (2007). The Italian versions of the Beck Anxiety Inventory and the Beck Depression Inventory-II: Psychometric properties and discriminant power. In Leading-edge psychological tests and testing research. (pp. 27–50). Nova Science Publishers. http://ovidsp.ovid.com/ovidweb.cgi?T=JS&PAGE=reference&D=psyc5&NEWS=N&AN=2007-13441-002

Spielberger, C. D., Gorsuch, R. L., & Lushene, R. E. (1970). The State-Trait Anxiety Inventory (STAI) Test Manual for Form X. ((tr. it.:). Consulting Psychologist Press.

Trakada, A., Nikolaidis, P. T., Economou, N. T., Kallianos, A., Nena, E., Steiropoulos, P., Knechtle, B., & Trakada, G. (2022). Comparison of sleep characteristics during the first and second period of restrictive measures due to COVID-19 pandemic in Greece. European Review for Medical and Pharmacological Sciences, 26(4), 1382–1387. https://doi.org/10.26355/eurrev_202202_28131

Ueda, M., Stickley, A., Sueki, H., & Matsubayashi, T. (2020). Mental health status of the general population in Japan during the COVID-19 pandemic. Psychiatry and Clinical Neurosciences, 74(9), 505–506. https://doi.org/10.1111/PCN.13105

Viselli, L., Salfi, F., D’atri, A., Amicucci, G., & Ferrara, M. (2021). Sleep quality, insomnia symptoms, and depressive symptomatology among italian university students before and during the covid-19 lockdown. International Journal of Environmental Research and Public Health, 18(24). https://doi.org/10.3390/ijerph182413346

Weissbourd, R., Batanova, M., Lovison, V., & Torres, E. (2020). Loneliness in America How the Pandemic Has Deepened an Epidemic of Loneliness and What We Can Do About It. In Harvard Graduate School of Education: Making Caring Common.

Wetherall, K., Cleare, S., McClelland, H., Melson, A. J., Niedzwiedz, C. L., O’Carroll, R. E., O’Connor, D. B., Platt, S., Scowcroft, E., Watson, B., Zortea, T., Ferguson, E., Robb, K. A., & O’Connor, R. C. (2022). Mental health and well-being during the second wave of COVID-19: longitudinal analyses of the UK COVID-19 Mental Health and Wellbeing study (UK COVID-MH). BJPsych Open, 8(4), 1–10. https://doi.org/10.1192/bjo.2022.58

